# *“We can’t develop standards for a thing which doesn’t belong to us”:* Assessment of infrastructure, behaviours and user satisfaction of Guardian Waiting Shelters for secondary level hospitals in southern Malawi

**DOI:** 10.1101/2023.11.02.23297991

**Authors:** Mindy Panulo, Jennifer Lamb, Kondwani Chidziwisano, Blessings White, Robert Dreibelbis, Tracy Morse

## Abstract

**Background:** Guardian Waiting Shelters (GWSs) are an essential component of the Malawi’s health care system as they serve as a temporary home for patient guardians while taking care of their relatives admitted to the hospital. Despite GWSs valuable role in healthcare settings, there have been few studies on the specific experiences, infrastructure, and conditions provided at these facilities. The study examines GWS management structures and conditions, as well as guardian satisfaction and perception of health risks related to GWS use.

**Methods:** In this cross-sectional, mixed-methods study, we assessed 12 GWSs from 12 districts in the southern region of Malawi. Qualitative data included interviews (n=149) and focus group discussions with patient guardians (n=72), as well as interviews with GWS caretakers (n=5), representatives from Hospital Management (n=12) and Hospital Advisory Committees (n=11). Quantitative data included structured assessments (n=12) of infrastructure present and used at GWSs. Descriptive statistics and qualitative thematic analysis were utilized for data analysis, and a problem tree analysis was used to triangulate and summarize the findings.

**Results:** 249 participants, including 221 being patient guardians, participated across the 12 GWSs. Each GWS had an average of 100 users daily, primarily middle-aged females (71%). There was a lack of clear and consistent management and responsibility for GWS operation and maintenance. GWS infrastructure conditions were poor, with inadequate functional sleeping rooms, insufficient access to water and sanitation facilities, and limited facilities for hygienic food preparation. Notably, 50% of the GWSs lacked water access, and a quarter had non-functional toilets. Overall, guardians felt unsafe and at risk of disease transmission when staying within GWS.

**Conclusion:** Study findings highlight lack of clear, consistent GWS ownership as a root cause of challenges in GWSs. Clear policy and operational standards must be established for effective management and smooth functioning of GWSs in Malawi.

## BACKGROUND

Sustainable Development Goal (SDG) 3.8 aims to achieve universal health coverage (1). However, Low- and Middle-Income Countries (LMICs) are struggling to achieve SDG 3 not only as a result of the recent competing demands of COVID-19 pandemic, but also the existing shortage of healthcare personnel, supplies and budget (2–6). As a result, hospitals in the sub-Saharan Africa region, including Malawi, rely on family members as patient’s carers (locally known as patient guardians) as a key component of health services.

In Malawi, guardians are typically accommodated in a dormitory type of housing known as the Guardian Waiting Shelters (GWSs) (7–9). GWSs are today an integral component of the health care system in Malawi, established to provide a safe and healthy environment for patient guardians to reside in while they attend to their relatives admitted to hospital. Duties of these guardians at hospitals typically involve activities such as bathing, cooking, feeding, monitoring of medications, and assisting with rehabilitation exercises for patients (2,9,10). Additionally, the physical presence of a family member during hospitalization has shown to offer psychosocial support to the patient, which ensures trust in the care the patient is receiving (11). In some circumstances, GWSs also serve as Maternity Waiting Homes (MWHs) for women with high-risk pregnancies or those from hard-to-reach areas as they await their expected date of delivery (11–14). The World Health Organisation (WHO) recommends provision of MWHs in healthcare settings to promote Maternal Child Health (MCH) (15). However, in Malawi, specifically designated MWHs have been reported in only 50% of the district hospitals (9).

Despite GWSs providing residence to patient guardians and maternity waiting mothers, previous research has focused on MWHs only, through the examination of their conditions, usage, quality of care, users experience and satisfaction (12,16–20). Evaluation studies in Malawi, Ethiopia and Zambia found that MWH users were dissatisfied with the sleeping and cooking space, availability of water, lack of privacy, poor sanitation, pests and congestion (12,18,21,22). In contrast, a qualitative thematic analysis recommended clean environments and safe structures as essential in effective functioning of the MWH (23). To date, there has been limited research to evaluate the status of the GWS model as an integral component of the general healthcare service delivery system in Malawi and similar contexts.

Thus, this study was conducted to 1) examine the GWS management structures and available infrastructure and services; 2) understand guardian satisfaction with the GWS structure and environment, and their perception of health risk from GWS use. Results from this study will inform service providers, local planners, and decision makers to improve the functionality of GWSs, and will examine the current and potential roles patient guardians can play in this.

## METHODOLOGY

### Study design and setting

This was a cross-sectional exploratory sequential mixed-methods design where structured quantitative observation data was used to inform subsequent qualitative data collection.

The study was conducted in twelve GWSs located across the twelve districts in the southern region of Malawi. These hospitals belong to the second tier of Malawi’s healthcare system services as referral centres for primary healthcare facilities. The hospitals offer outpatient and inpatient services, surgical procedures such as caesarean sections, and other emergency life-saving surgeries.

### Study Population

We recruited patient guardians who were using the GWS, members of the Hospital Advisory Committee (HAC), members of the hospital management committee and a GWS caretaker where available. A guardian was a family member or designated carer of the patient seeking health care. Members of the Hospital Advisory Committee (HAC) were community representatives who act as mediators between the community members and healthcare workers. They also ensured that there was accountability of medicine at the hospital, the hospital grounds and services were respected and functional, and delivered hygiene talks to patient guardians. The Hospital Management Committee (HMC) was tasked with the daily management of the hospital, and included the District Medical Officer, District Environmental Health Officer, Hospital Administrator etc. The HMC also played a pivotal role within their community e.g., during immunisation campaigns such as polio, and during the pandemic they were active to counter misconceptions about COVID-19. GWS caretakers were personnel employed by the hospital. Their roles included sweeping the public yard area of the GWS, cleaning toilets, solid waste management, ensuring there was water onsite at the GWS and reminding patient guardians to take care of the sleeping, cooking areas and water, sanitation, and hygiene (WASH) infrastructure.

### Sampling and sample size

The study targeted ten district (public) hospitals and two Christian Health Association (CHAM) (private) hospitals in the southern region of Malawi (Figure 1).

**Figure 1:**
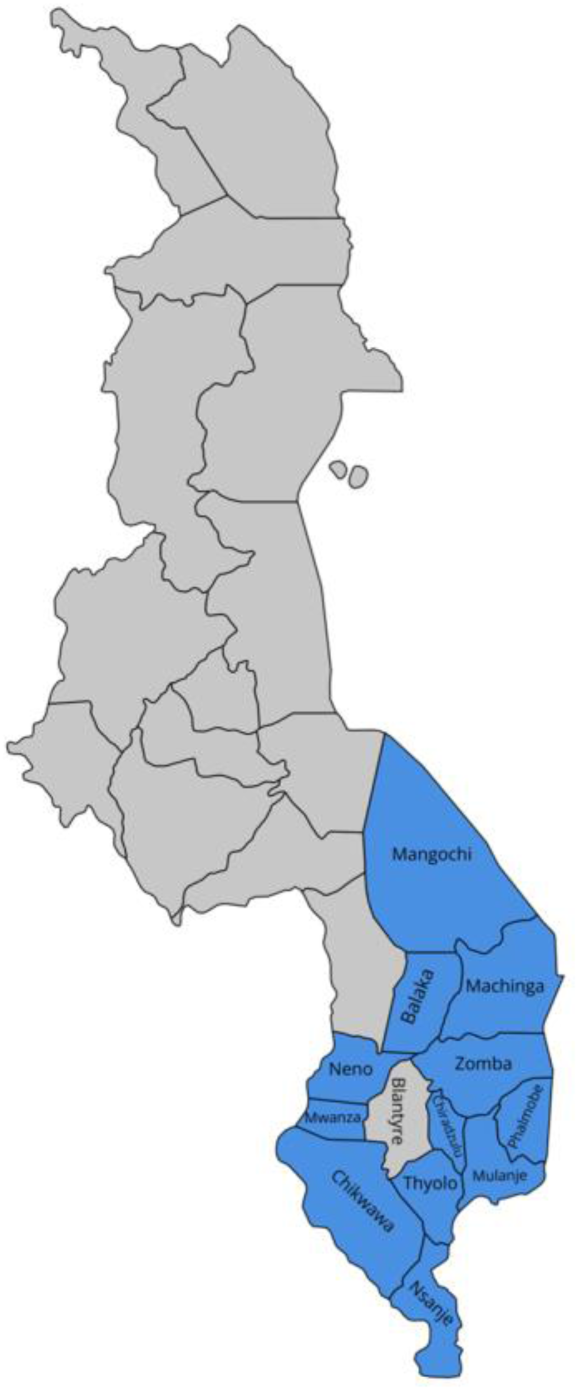
Map of Malawi depicting the Districts of the Southern Region included in the study (Blantyre not included in the sample as it does not have secondary level hospital facility and was therefore excluded)

From each hospital, we recruited 1 HAC chairperson, 1 GWS caretaker, 1 Hospital Management Committee member and six patient guardians for in-depth interviews (IDIs). Two focus group discussions (FGDs) were also conducted at each hospital (1 male; 1 female) to fully understand the opportunities and challenges surrounding GWSs. The study recruited patient guardians who had used the GWS for more than four days to ensure they were familiar with the hospital and GWS setting.

### Data collection

Data was collected from 20^th^ December 2021 to 25^th^ March 2022 by three experienced research assistants who were fluent with the local language (*Chichewa)*. These research assistants were trained for four days including pre-testing of the data collection tools prior to data collection. The study applied various methods of data collection to ensure a comprehensive understanding of each GWS setting:

#### Observation checklist

An observation checklist was used to assess availability and condition of infrastructure at the GWS, which included sleeping rooms, access to safe water, hygiene, and sanitary facilities. These included bathing areas, toilets, water points, handwashing facilities, laundry areas and cooking places. Observations provided insights for further exploration during the subsequent interviews and focus group discussions.

#### Qualitative data

In-depth interviews (IDIs) were conducted with patient guardians to understand their daily life at the GWS, including hygiene practices, access to water and sanitary facilities, perception of safety and disease risk while staying at the GWS. These also provided a snapshot of the role of the patient guardians in health care utilisation for their family member who is admitted at the hospital. IDIs with HMC and HAC representatives provided an understanding of the ownership, governance structures, and daily running of the GWS. IDIs with GWS caretakers generated information on daily management of the GWS. FGDs with patient guardians were conducted to understand social and community regulated practices in each locality based upon shared common values and beliefs, which could not be uncovered using IDIs.

IDIs lasted 45 to 60 minutes while FGDs were conducted for 60 to 90 minutes in the local language.

### Data analysis

#### Quantitative data

Quantitative data from checklist observations were collected through Kobo Collect software platform (24) and later exported to Microsoft Excel 2016 (Microsoft Corporation, Redmond, WA, USA). Descriptive statistics were used to summarize the collected data and categorical variables were summarized using frequencies.

#### Qualitative data

All interviews (IDI and FGD) were audio recorded which helped to manually generate preliminary transcripts from the interviews. Transcriptions were read and verified by four research team members before coding them based on deductive study themes of management structures, perception of health risk among GWS users and satisfaction with the GWS structure and environment. The analysis then enabled inclusion of inductive themes.

#### Problem analysis

Taking into account the findings of the assessment across all 12 districts, the research team examined the qualitative and quantitative data using a problem tree analysis to map out the anatomy of the cause-and-effect relationships seen at the GWS (25).

### Ethical consideration

This study was approved by the National Health Science Research Committee of Malawi (P No. 21/11/2822) and London School of Hygiene and Tropical Medicine Ethics Committee, UK (Ref 2655). All interviewed respondents provided written informed consent for participation in the study, and no names were mentioned or recorded during the FGDs and interviews.

## Results

### Socio-demographic characteristics of respondents

In total, 249 respondents were involved in the study, with 221 participants being patient guardians. Seventy-two patient guardians were recruited for IDIs while 149 patient guardians were involved in FGDs. Only five GWSs had caretakers who participated in the study (Table 1). Eleven management committee members and 12 HAC members participated through IDIs.

**Table 1:** Study participants.

| GWS name | Affiliation | HAC<br>chairpersons<br>(IDI) | Management<br>committee<br>member (IDI) | GWS<br>caretaker<br>(IDI) | Patient guardians |  |
| --- | --- | --- | --- | --- | --- | --- |
|  |  |  |  |  | FGDs | IDIs |
| Chiradzulu District Hospital GWS | Public HCF | 1 | 1 | - | 6 | 6 |
| Thyolo District Hospital GWS | Public HCF | 1 | 1 | 1 | 14 | 6 |
| Mulanje District Hospital GWS | Public HCF | 1 | 1 | 1 | 7 | 6 |
| Zomba St. Lukes GWS | Private HCF | 1 | - | 1 | 9 | 6 |
| Machinga District Hospital GWS | Public HCF | 1 | 1 | - | 16 | 6 |
| Mangochi District Hospital GWS | Public HCF | 1 | 1 | 1 | 14 | 6 |
| Chikwawa District Hospital GWS | Public HCF | 1 | 1 | - | 14 | 6 |
| Nsanje District Hospital GWS | Public HCF | 1 | 1 | - | 12 | 6 |
| Balaka District Hospital GWS | Public HCF | 1 | 1 | - | 14 | 6 |
| Mwanza District Hospital GWS | Public HCF | 1 | 1 | - | 16 | 6 |
| Phalombe Holy Family GWS | Private HCF | 1 | 1 | 1 | 13 | 6 |
| Neno District Hospital GWS | Public HCF | 1 | 1 | - | 14 | 6 |
| **Total** |  | 12 | 11 | 5 | 149 | 72 |

Across all 221 guardians who participated in the study, the average age was 40 years with 43% of them being a patient guardian to antenatal waiting mothers (43%) (Appendix 1). Seventy-nine percent of the patient guardians depended on public transport to get to the hospital/GWS and on average they had stayed at the GWS for 19 days prior to data collection.

**APPENDIX 1:** Characteristics of the patient guardians (n=221)

| Variable | Percentage<br>(n=221) |
| --- | --- |
| **Guardian's relationship to patient** |  |
| Brother | 13.55 |
| Daughter | 39.25 |
| Daughter in law | 4.21 |
| Mother | 3.74 |
| Sister | 13.08 |
| Son | 9.35 |
| Others (Wife, Grandchild, sister-in-law, mother-in-law, uncle, and father) | 16.82 |
| **Level of education** |  |
| Never attended | 23.53 |
| Primary | 62.44 |
| Secondary | 13.12 |
| Tertiary | 0.91 |

### Ownership, and stakeholders responsible for conditions and management of the GWS

#### Ownership

Two of the GWSs were serving privately owned faith-based (Christian Health Association of Malawi) healthcare facilities and were managed by the environmental health department through the hospital administrator. The remaining 10 GWSs were affiliated with government-owned public hospitals. There was significant variation in management and oversight of GWSs at government facilities (Table 2). Several stakeholders including the Members of Parliament, District Councils, Hospitals, and the wider community were identified as key stakeholders in the daily operation and maintenance and routine monitoring of the GWS. Interviews revealed a lack of clarity on who was responsible for GWS across multiple stakeholders. In several cases, respondents said that the GWSs were the responsibility of the hospital management since the GWSs were usually built on or near hospital grounds, and served people who were using the hospital. In other interviews, the hospital management and HAC members explained that the GWS belonged to the district council or the community.

**Table 2:**
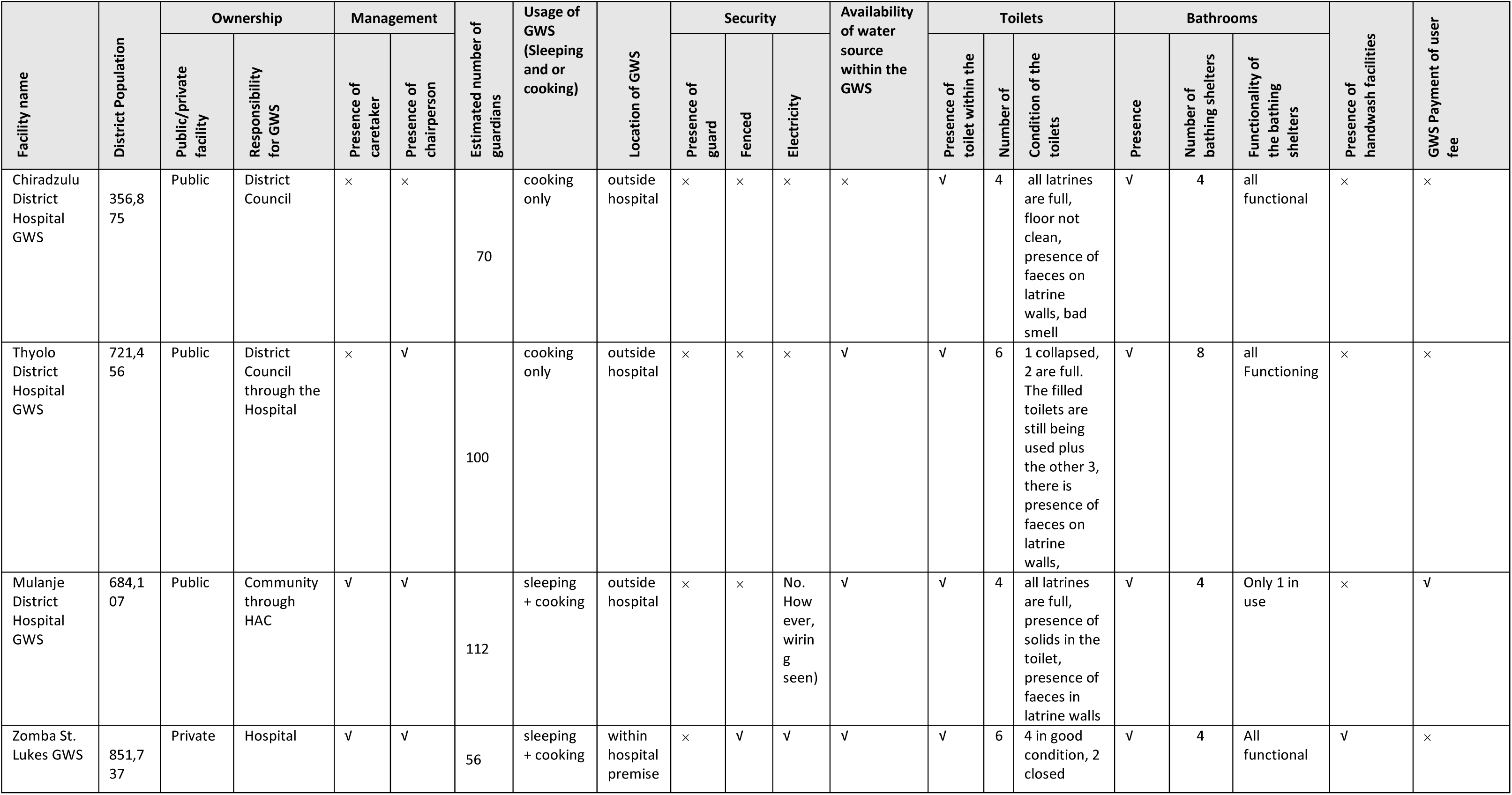
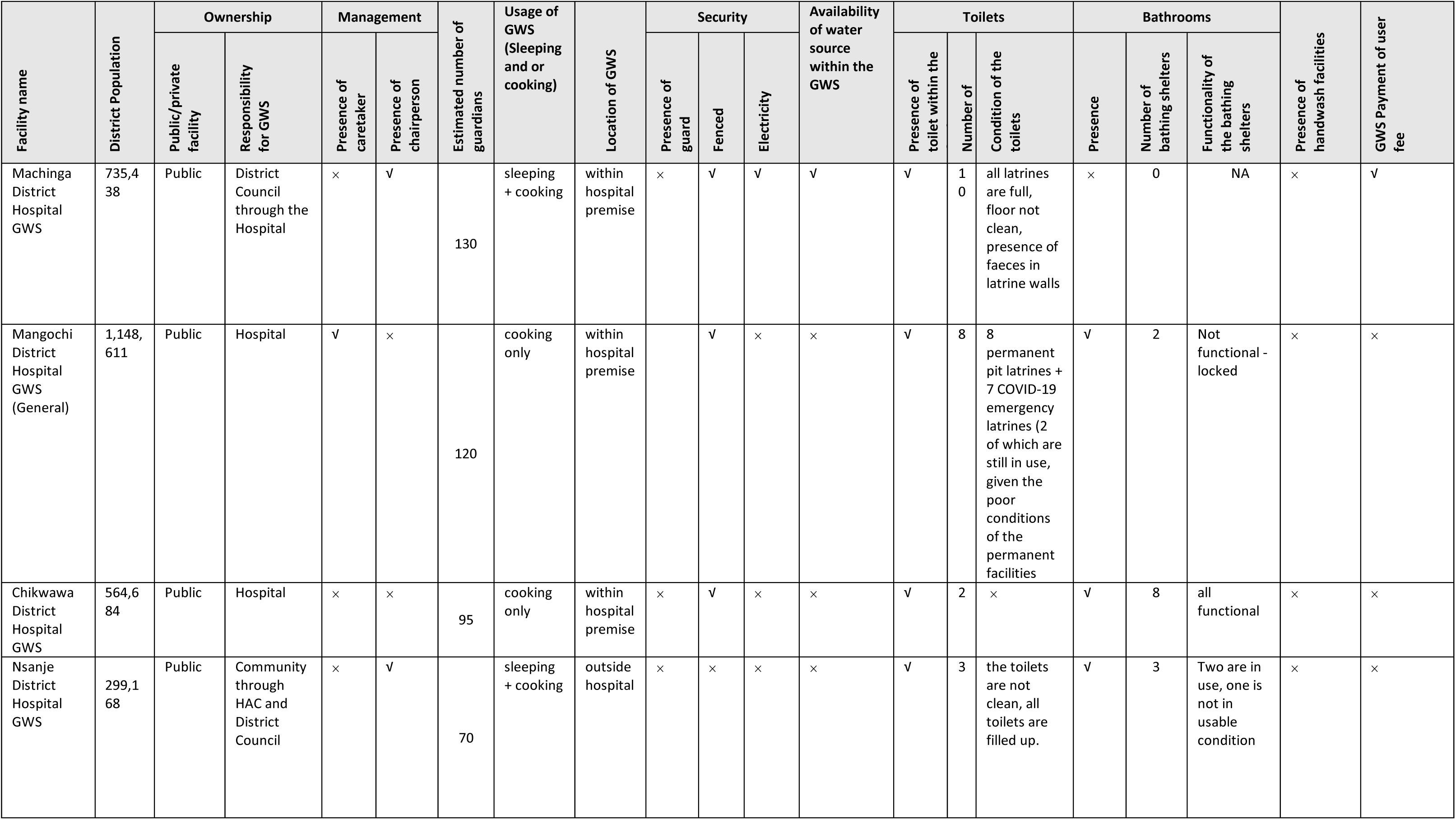
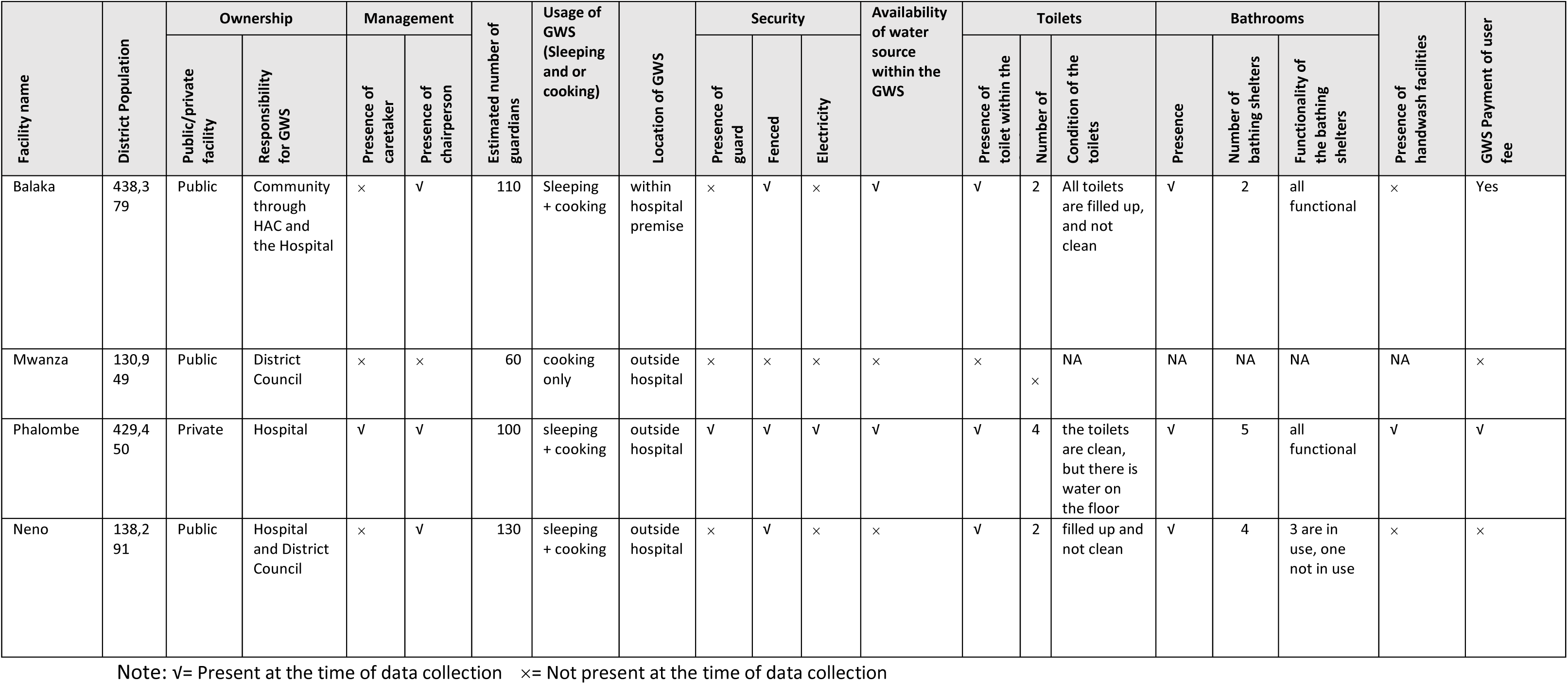
GWS Characteristics for twelve southern region health facilities.

#### Budget

Only GWSs serving private owned healthcare facilities had an allocated budget. For public facilities, respondents reported using limited resources that were meant for patient wards, such as gloves and bleach for cleaning and disinfection to take care of the GWS.

> “No. How can we budget for a thing which is not owned by us, it belongs to the district council. Sometimes as a hospital we just help because the GWS is near us’’ (IDI, Hospital management representative)

#### Stakeholders responsible for conditions and management

Five GWSs had a paid caretaker (two from private and three from public facilities). The caretaker was responsible for cleaning the bathrooms, toilets, and the surrounding environment. At nine GWSs, a chairperson for each sleeping room was appointed from among the guardians staying in the facility (Table 3). The GWS chairperson was responsible for maintaining order at the GWS, solving disputes among patient guardians, allocating sleeping space to new guardians, and organising cleaning of the GWS. GWSs with a caretaker had generally better hygienic conditions than those without caretakers while there were no noticeable improvements in hygienic conditions among facilities with and without chairpersons. GWS chairpersons reported that they had limited authority as volunteers and other guardians did not respect them.

> “It is difficult to maintain cleanliness here at the GWS because each time I try to organise my fellow patient guardians to clean the shelter, they always talk ill saying this is not my home, just mind your own business’’ (IDI, GWS Chairperson).

Respondents acknowledged that GWSs suffer from the lack of standards and policies to ensure there is a common strategy to design, deliver, operate, maintain, and monitor the facilities. This issue was further exacerbated by the absence of clarity of ownership and governance of GWSs at the district and national level.

> ‘’We can’t develop standards for a thing which doesn’t belong to us. The first thing to be established is ownership for that facility, from there, issues of standards and monitoring can be resolved’’ (IDI, DEHO)

Some respondents highlighted the absence of an Infection Prevention and Control (IPC) policy targeting GWSs as *“it is associated as being low risk” (IDI, DEHO)* and beyond the scope of existing guidelines focusing on medical facilities. Meanwhile, one respondent challenged that an IPC policy should also focus on the GWSs with the following reason.

> ‘’That area can be an important source of diseases. For example, people are sleeping on floors, so in terms of scabies, the diseases can spread very fast” (KII, DEHO).

Several HAC respondents raised concerns about tensions between HAC members and guardians. Guardians often disregarded requests from HAC members to take specific actions to improve conditions in GWSs (e.g., keeping latrines, cooking, and sleeping rooms clean and tidy). HAC members suggested this was linked to the lack of any visible identification – such as an identification (ID) or uniform – that signified their official role within the health system. Caretakers were also responsible for encouraging hygienic use and maintenance of GWS infrastructure.

### Guardian Waiting Shelters use and characteristics

During the time of the study, an average of 100 people were reported to be utilising each GWS on a daily basis. For the five facilities that had no specific accommodation for the maternity waiting mothers (Maternity Waiting Shelters), GWSs were used to also accommodate both maternity waiting mothers and patient guardians.

Generally, staying at the GWS was free of charge, however, four GWSs collected fees (1 private and 3 public GWS) of approximately USD0.2 per patient guardian. Collection of the fees was locally arranged by the GWS caretaker or GWS chairperson and collection frequency varied from daily to only collected as the need arose. GWS chairpersons reported that the money was used for buying cleaning materials (e.g., hoes, brushes, brooms, and mops), and lighting materials (e.g., candles and torches).

Seven of the GWSs had sleeping rooms used by the patient guardians for sleeping (Table 2)(Figure 2). In the rest of the five GWSs, three were not used by the patient guardians due to poor conditions (i.e., no doors, windows and partly roofed), while two of GWS had no sleeping rooms. In these cases, the patient guardians reported sleeping in patient wards, hospital verandas and corridors (Table 2). None of the GWSs had a supply of basic needs and amenities such as mosquito nets, bedding, lockable sleeping room doors or a secure place to store their belongings. Ten GWSs, all serving public facilities, lacked electric lighting (Table 2). GWS serving private owned healthcare facilities had better amenities such as electricity, security guards, and perimeter fencing around the GWS (Table 2).

**Figure 2:**
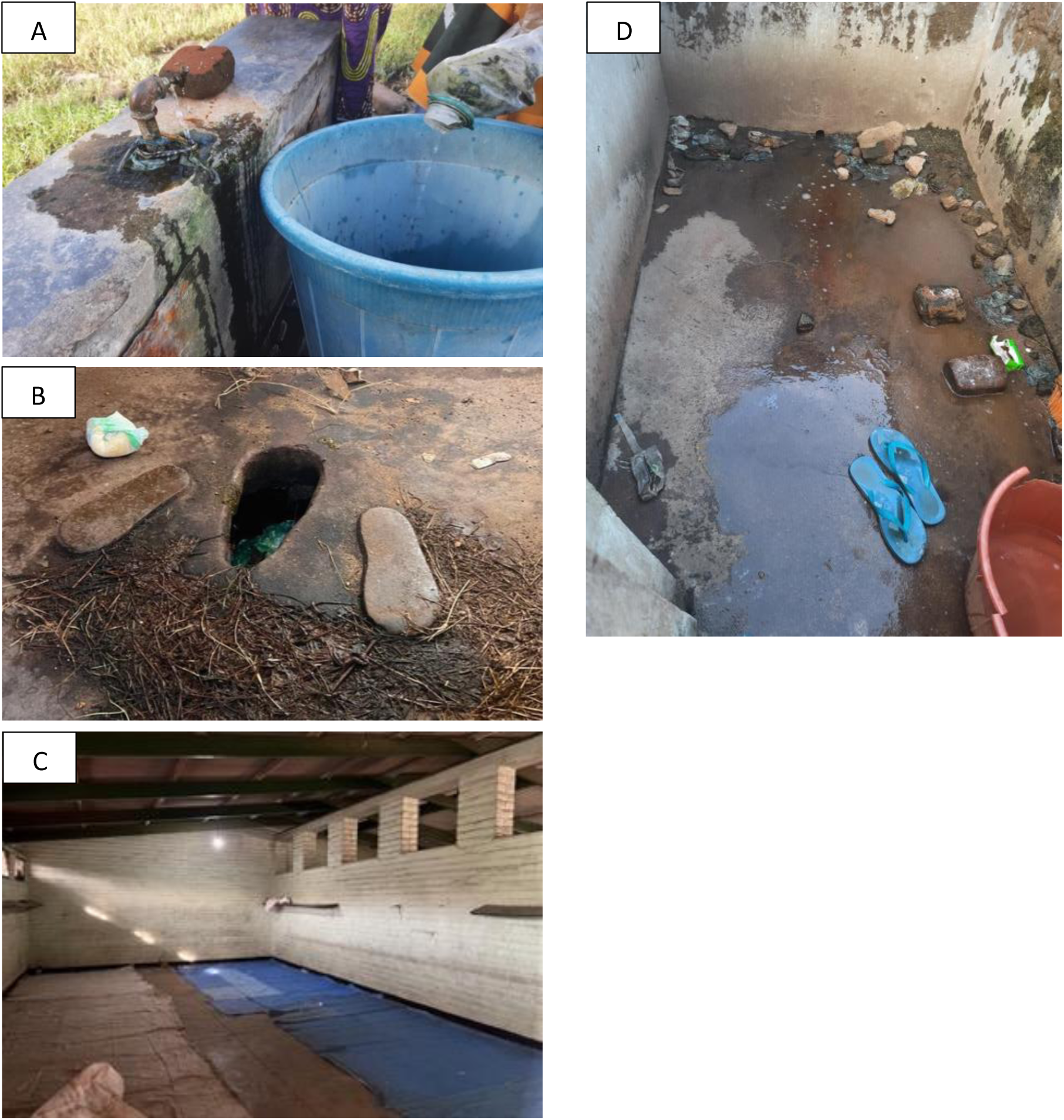
GWS characteristics observed during data collection A: Broken water tap located at GWS B: GWS pit latrine which is full, with indiscriminate disposal of diapers C: Standard sleeping room for GWS D: GWS bathing shelter used by both make and female patient guardians

Six out of the twelve GWSs had a functional onsite water supply from a safe source (standpipes and boreholes) within the GWS, while others accessed water either from community, school and hospital water points that included boreholes and standpipes (Table 3). Missing or broken taps were commonly observed at GWSs (Figure 2). Respondents reported lack of timely and correct payment of pre-paid water, pumps being broken and sporadic electricity limiting water availability. Eleven of the GWSs had toilets (Table 2), which included the two GWSs serving private owned healthcare facilities which had both water closet toilets and pit latrines. The remaining nine GWSs serving public owned healthcare facilities had only pit latrines, of which seven were almost full and faeces, used nappies, and menstrual hygiene materials were observed on the floor (Table 2). Three GWSs had evidence of open defaecation.

> “Sometimes I prefer to go to the bush than using the latrines which are very unhygienic’’ (Female IDI participant).

Hygiene conditions were generally better in GWSs at private owned healthcare facilities compared to those serving public owned healthcare facilities. Handwashing facilities (manually filled 20-litre buckets with taps) were observed only in the GWSs at private owned healthcare facilities (Table 2). However, soap (liquid or bar) was not observed at any HWF. The majority of GWSs (n=11) had adequate facilities for guardians to bathe (bathrooms). However, GWSs at public owned healthcare facilities were observed to be in poor condition (i.e., poor drainage, no privacy and were left uncleaned)(Figure 2). Due to unhygienic conditions of the toilets, guardians reported feeling disgusted by latrine conditions and resorted to using bathing areas as toilets and or opted for open defaecation. Only one GWS had information education and communication (IEC) materials (poster) displayed within the GWS to promote handwashing with soap.

Five GWSs had rubbish pits which were found to be full of waste from the GWS, and often located close to the cooking areas. Market sellers nearby the GWSs were also reported to use the GWS infrastructure – specifically toilets and rubbish pits. Additionally, people with developmental disabilities were also reported and observed staying in five of the GWS, including using the sleeping and cooking places.

Eight of the GWS reported to offer health and hygiene talks to the patient guardians either on a daily, weekly, or ad-hoc basis (e.g., during outbreaks). These talks were conducted by the GWS caretaker, guards, HAC members, or health personnel, and were confirmed by the patient guardians. Talks contained various messages covering sanitation (solid waste management, use of latrines), hand hygiene and potential risks and health concerns of diarrhoea, cholera, COVID-19, scabies, malaria, cervical cancer, Tuberculosis, and HIV.

### Guardian and Caretaker Experiences using Guardian Waiting Shelters

There were mixed receptions to payment among guardians, some felt it was a good idea since this money was used to improve their wellbeing and overall GWS conditions, while others did not appreciate this approach as they already had financial challenges to take care of their patients and had the expectation that the hospital management should cover such costs for the GWSs.

#### Water, sanitation, and hygiene (WASH)

All patient guardians appreciated the presence of the GWS within the hospital and reported using the GWS as their home as it offered them a place to cook, bath and some to sleep. At better maintained facilities, guardians reported being able to practise selected hygiene behaviours while at the GWS such as bathing, sweeping and mopping sleeping and cooking areas, hand washing before eating, washing clothes, washing kitchen utensils, washing food before cooking, or covering food and water, and disposing or reheating left-over food. Motivation to practise such behaviours were reported as preventing disease, a continuation of what they practised at home, and having more free time while at the GWS. Additionally, some patient guardians reported that since they found the GWS already clean, they were motivated to ensure they maintained these standards.

However, where infrastructure was limited or poor, patient guardians reported poor hygiene practices. For example, due to lack of laundry spaces, patient guardians washed soiled clothing including bedding at drinking water collection points, which discouraged others from using such water points and resulted in them collecting water from alternative sources located outside the GWS premise. It was further reported that female patient guardians were leaving menstrual hygiene materials in the bathing shelters, as they had no appropriate means for disposal or management. Since most of the GWS were utilised by women, shelters for bathing were not segregated by gender and one male IDI participant commented:

> “Sometimes I fail to bath here at the GWS because it is shameful for me to find menstrual hygiene materials in the bathrooms’’ (IDI, Male guardian).

#### Cooking and food hygiene

Cooking meals for patients was considered one of the primary responsibilities of the patient guardians. Guardians cooked from various locations within the GWS, e.g. on the ground of the GWS compound area, in old sleeping rooms or in designated kitchens. Food preparation was generally considered challenging by all guardians. Guardians reported that the cooking rooms were inadequate and often lacked ventilation, especially considering that main source of fuel was biomass. The lack of handwashing facilities in the cooking spaces was reported to lead to inadequate or no handwashing during cooking. Limited kitchen utensils availability (e.g., dish racks and plates) was further reported to compromise food hygiene practices. Patient guardians felt that the lack of WASH facilities compromised their ability to practice food hygiene which may subsequently affect both guardians and patients.

> “Due to lack of proper cooking place, we cook in those old sleeping rooms which are small and poorly ventilated. We fear this may cause cough and other respiratory diseases’’ (Female FGD participant).

#### Perceptions of public health risk

Patient guardians perceived that the poor environmental health conditions at the GWS, such as overfilled waste disposal sites, poor ventilation, limited access to safe water, sanitation, and hygiene (WASH) infrastructure and poor cleaning standards, exposed them to specific health risk. Specifically, the patient guardians felt vulnerable to the transmission of cholera, diarrhoea, skin diseases, COVID 19, malaria, cough, colds, Tuberculosis (TB), and pneumonia when staying at the GWS.

Patient guardians from public owned health facilities also highlighted safety, security, privacy, and well-being concerns. This was related to the location of the GWS, which were often far from the hospital, congested sleeping rooms (three people per square metre) (Figure 2), broken windows, poor lighting, lack of locks on the sleeping room doors, and no means to secure their belongings in lockers.

> “This GWS is isolated, with no fence nor electricity such that we are never safe as women” (Female FGD participant).

Staff responsible for delivering health training reported that all support they received focused on roles and responsibilities of GWS users and had limited technical training on topics they were required to cover in health and hygiene talks. Instead, they used and applied their own knowledge acquired from other roles when delivering the health and hygiene talks at the GWS. Additionally, despite healthcare facility management reporting that the talks were delivered regularly, guardians reported that these talks were infrequent:

“The caretaker or chairperson call us at one place to communicate on a particular issue, but this only happens occasionally…. since I came here a week ago, we have been briefed on hygiene issues once” (IDI, female guardian).

#### Social relationships

Guardians were very dependent on financial support during the time they spent at the facility – usually receiving money, food and firewood from friends and neighbours. Many reported that financial support was insufficient, leading them to seek piece work in the surrounding area, often resulting in tensions with local vendors or laborers. Additionally, patient guardians also reported fetching and selling firewood to other patient guardians at GWS to have money. Quality of the relationships between patient guardians varied across the GWSs. In some instances, patient guardians indicated good relationships in terms of sharing food and firewood and escorting one another during the night from the GWS to the hospital. On the other hand, some patient guardians complained of theft between patient guardians, taking items such as cooking utensils, clothes, and food.

> “I remember I came to the hospital unplanned as my patient had an emergency condition. But when I arrived here at the GWS, my fellow patient guardians gave me food for my patient’’ (IDI, Male guardian).

#### Accountability

Channels by which patient guardians could engage with those in charge of the GWSs were unclear. This was compounded in many cases by the lack of clarity, even at management level, of where responsibility for this facility lay. Even when reporting mechanisms were clear, patient guardians reported that they were not comfortable reporting challenges, as they felt doing so would compromise the quality of care given to their patients. In some instances where patient guardians were nominated as the GWS chairperson there was also a belief that the associated patient would not get better and not be discharged within a short period of time. Such a myth therefore potentially discouraged patient guardians volunteering for such a role when staying at the GWS.

### Problem analysis

Based on understanding from both interviews and literature that patient guardians are an integral yet neglected part of the health care system in Malawi. We used a problem tree analysis to triangulate the results and summarise the environmental, social, and economic challenges for patient guardians to effectively utilise GWSs in their current state (Figure 3).

**Figure 3:**
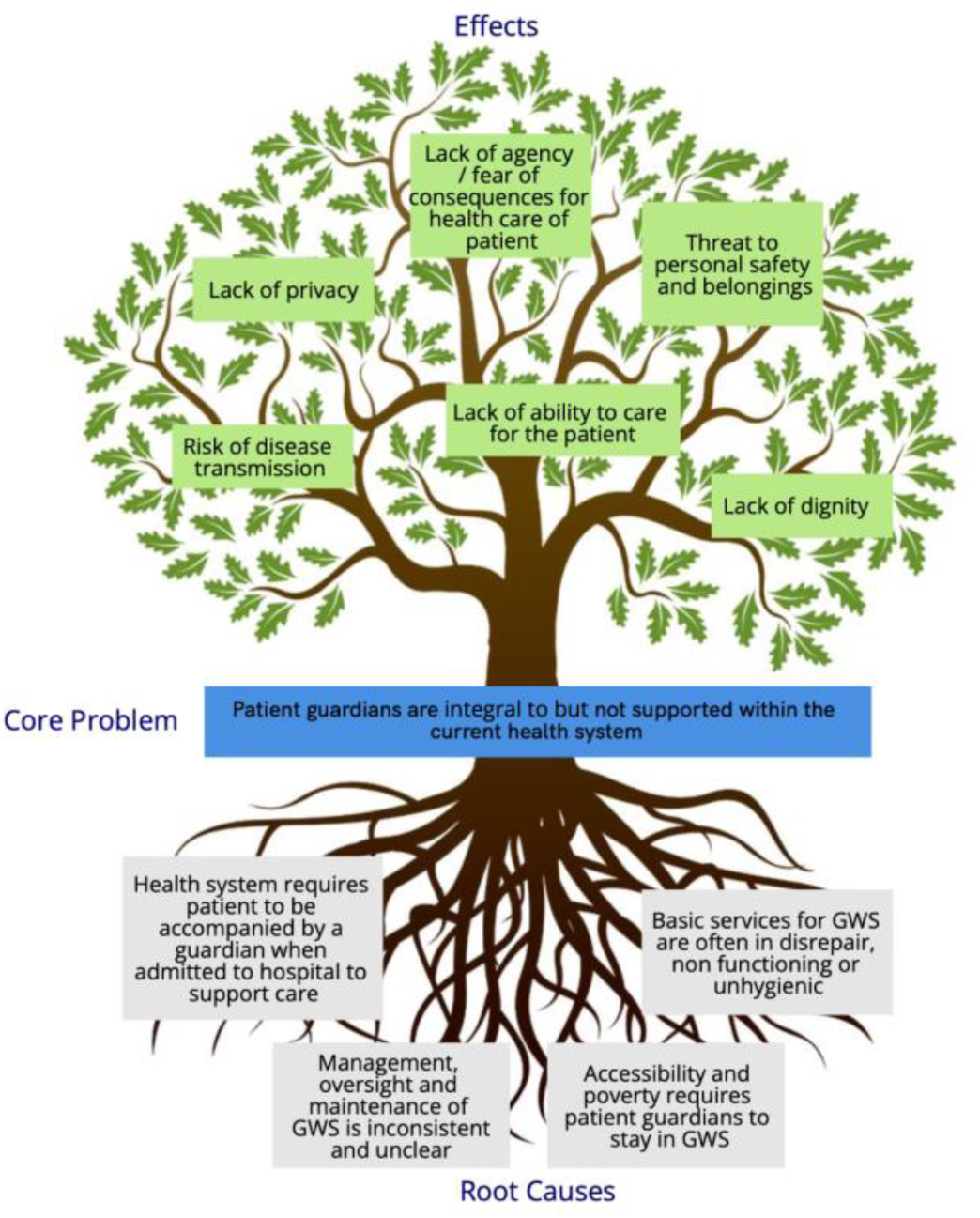
Problem tree analysis summarising findings of GWS assessment

## Discussion

Most sub-Saharan African countries including Malawi adopted the patient guardian model where the patient guardians are responsible for providing informal supportive care to their relatives who have been admitted at the hospital (26,27). Over time, these patient guardians have proven to be useful assets in the health system as they reduce workload among overburdened healthcare workers and improve the quality of healthcare services which patients receive while in hospital, albeit the informal nature of their role continues to challenge the system (2,10,28–30). However, little is known about the conditions in which these patient guardians are residing. The study established that the GWSs are utilised by not only patient guardians but also serve as maternity waiting homes (MWHs) and are used daily by local vendors and members of the community with disabilities who have no formal support service.

It is acknowledged that many of the factors identified in this study are underpinned by the challenges faced in least developed countries such as Malawi, not least poverty which continues to be the main driver of these. Therefore, this study has considered these underlying factors, and identified three key areas which could realistically be addressed to improve the patient guardian experience, and therefore patient care, as well as reducing the risk of GWS acting a vehicle for infection transmission.

### (1) Full integration of GWSs into the health care system

The lack of clear ownership and accountability for the GWS was a consistent root cause of other challenges facing effective management and maintenance of the facilities for patient guardians. This was particularly pronounced at public health facilities, where the lack of clear ownership, budget, and accountability systems to ensure effective conditions and management, make it near impossible to manage access and maintain better standards for the patient guardians. Where improved systems were in place, such as employed caretakers or privately manged facilities, GWSs were able to maintain more hygienic and acceptable standards. Clarifying roles and responsibilities for management and maintenance of facilities has also been shown to be effective for the management of MWH where policies, management roles and finances are incorporated in the MWH model (31), and there is a clear recognition that health facilities are central to providing this support (32). Placing GWSs under direct and clear hospital management would also offer a strategic opportunity to formalise existing supporting structures such as the HAC who already have an informal role in GWS operations and have been found to be an effective model for oversight and management of health facilities in other contexts (33). Clearer management would also enable the role of GWS chairpersons to be re-evaluated, providing an opportunity to serve as a conduit for patient guardian communications and support the development of social capital in the group, rather than oversight of the facility for which they do not have the authority and are not compensated. More formal management would also enable clearer and realistic training for caretakers and chairpersons on technical subjects such as mitigating the risk of diarrhoeal and respiratory infections and providing them with appropriate acknowledgement of their role (e.g., ID, uniform).

### (2) Development of clear standards for GWSs which encompass minimum requirements for infrastructure, services, and security

There are currently no documented standards for patient guardians shelters in Malawi, or similar settings, which outline minimum infrastructural requirements for these settings. Although all GWSs in this study had access to safe water, some collected the water from outside the GWS premise which could compromise the water quality and quantity and limit adequate hygiene behaviours (34) (35,36). Lack of hygienic sanitation facilities also drove some guardians to practice open defaecation in the immediate area, further eroding sanitary conditions of the GWSs and introducing additional disease risks. This study highlights the complex repercussions of poor infrastructure and services in these settings. Not surprisingly, the risk of disease transmission, as a result of poor WASH facilities, is high not only between patient guardians within the GWS, but also between the hospital and the GWS residents as people move between wards and overcrowded, poorly ventilated and unclean accommodation (37). However, it is also clear that the poor infrastructure and services have an additional impact on safety, security, dignity, and mental wellbeing of patient guardians. All of these reduce their ability to look after those in their care effectively, while addressing the additional stresses and burdens facing them around income and provision of support (2,38,39). With this in mind, a minimum standard for GWS infrastructure and services should be developed, as has been undertaken for other areas of the health service (40,41). The provision of these standards will support effective management and accountability, and provide the services needed for the third key area of integrating IPC.

### (3) Integration of GWS into hospital IPC systems

As reported elsewhere, the presence of guidelines for managing MWH in Malawi have contributed to smooth running and improved IPC of such premises (40,41). Therefore the development and integration of specific guidelines for the GWSs would be beneficial in improving the welfare of patient guardians while staying in the GWS. Guidelines should seek to encompass the GWS into the whole system of the health facility, taking advantage of readily available resources, and incorporating already existing IPC policy and guidelines for Malawi (40–42) to provide consistency of approach and a sense of ownership. However, they must also take into consideration the education and literacy levels of caretakers, HAC members and patient guardians, and as such an illustrative and behaviour centred approach to guideline development is essential if they are to be effective. Guidelines should also include the regular provision of health promotion activities which are delivered by skilled and competent personnel.

The study established mixed levels of social capital among the patient guardians. For those who indicated to have good relations with other fellow patient guardians, they expressed a better experience while staying at the GWS with a sense of belonging and support. High levels of social capital in a community has been proven to promote participation of community members on various development activities and improves their coping mechanisms (43–45). It is anticipated that if the standards and systems within the GWSs were to improve, this would lead to a similar improvement in social capital within the patient guardian population. With the observed limited resources at the GWSs, promoting social capital among patient guardians is vital in both improving and maintaining the conditions of the GWSs.

## Limitations

Some data from this study was based on self-reported data which is prone to bias as the participants may not recall all aspects of interest and may provide information based on what the researcher wants to hear. Where possible self-reported data has been validated through observation. GWSs in Malawi are present in both secondary and tertiary hospital settings; however, conducting the study in selected secondary level hospitals limits the generalizability of the present findings to other district and third tier hospitals in Malawi. Further, GWS management may have additional stakeholders at central government level to those recruited in this study; thus, future similar studies should consider targeting IPC and health facility infrastructure managers at Ministry of Health headquarters. Nevertheless, this study provides a better platform to understand the GWS context in Malawian hospitals for the subsequent design of interventions to improve the welfare of patient guardians.

## Conclusion and recommendations

GWSs remain integral to health care delivery in Malawi but are not effectively supported in current systems. The findings of this study call for the need for clear GWS management and ownership to facilitate improved welfare for the patient guardians, who plays an important role in health service delivery in Malawi. Such ownership should ensure that GWSs have dedicated budgets and staff. Importantly, much as specific guidelines for GWSs are essential, the need for the GWS management to be integrated into existing hospital IPC policy and guidelines cannot be overemphasized. Further, GWSs are potential touch points for health promotion among patient guardians and indirectly community members. Relatedly, future interventions aimed at improving patient guardian welfare in the GWSs should incorporate social capital strengthening. These study findings provide a basis from which a more detailed understanding of the GWS context in Malawi can be built.

## Acknowledgments

A special thanks to all District commissioners, all health facility administrators, District Health Officers, District Environmental Health Officers, and District Nursing officers for assisting us during the data collection. Additionally, we would like to thank all GWS caretakers, Health Advisory committee members and patient guardians who participated in the study. Lastly, we would like to thank Mr Stevie Amos who supported data collection and management.

## Data Availability Statement

All data referred to within this study available in a repository upon acceptance.

## Author Contributions

Conceptualization, M.P.,J.L., K.C. R.D., and T.M.; data curation, M.P.,J.L., K.C. R.D., and T.M; formal analysis, M.P., J.L., and B.W., funding acquisition, R.D.; methodology, M.P., J.L., K.C., R.D., and T.M.; resources, R.D.; software, M.P., and J.L.; validation, M.P.,J.L., K.C. R.D., and T.M.; visualization, M.P., J.L., K.C. and T.M.; writing—original draft, M.P., J.L., and T.M.; writing—review and editing, M.P.,J.L.,B.W., K.C. R.D., and T.M.;. All authors have read and agreed to the published version of the manuscript.

